# Natural History of Self-reported Symptoms Following SARS-CoV-2 Infection: A Target Trial Emulation in a Prospective Community-Based Cohort

**DOI:** 10.1101/2025.11.01.25339316

**Authors:** Yanhan Shen, Zach Shahn, McKaylee M. Robertson, Kelly Gebo, Denis Nash, CHASING COVID Cohort Study Team

## Abstract

**Background:** The natural history of symptoms after SARS-CoV-2 infection remains uncertain because many studies are non-representative, ignore background symptom prevalence, lack longitudinal tracking, and omit appropriate controls. Using a prospective, community-based cohort with repeated symptom measures, we estimated post-infection risks of long COVID symptoms versus contemporaneous uninfected controls.

**Methods:** We analyzed the CHASING COVID Cohort, a U.S. longitudinal study with surveys and serology (March 2020–December 2023). Infection status (January 2021–December 2022) was determined from self-reported PCR/antigen results, serology, or CSTE probable criteria. We emulated 24 monthly target trials comparing individuals newly infected at time zero with those remaining uninfected. Outcomes were new-onset long-COVID symptoms not reported pre-infection, assessed overall and within three clusters (neurological, autonomic, exercise intolerance) at 4–8 and 9–12 months post-infection. Inverse probability of treatment and censoring weights adjusted for confounding and informative loss to follow-up.

**Results:** The analysis included 1,055 infected and 52,310 uninfected person-trials. At 4–8 months, the adjusted risk of any long-COVID symptom was 22.6% (95% CI, 20.5–24.8) among infected versus 11.3% (11.1–11.5) among uninfected (adjusted risk difference [aRD], 11.3% [9.2–13.5]; adjusted risk ratio [aRR], 2.01 [1.81–2.20]). At 9–12 months, risks were 19.2% (17.0–21.3) vs 12.4% (12.2–12.7) (aRD, 6.7% [4.6–8.9]; aRR, 1.54 [1.37–1.72]). Across all three clusters, infected participants had consistently higher risks at both intervals.

**Conclusions:** SARS-CoV-2 infection was associated with elevated risk of new-onset long-COVID symptoms persisting up to 12 months. Using a national community-based cohort, contemporaneous uninfected controls, and target-trial emulation clarifies the burden attributable to infection and supports ongoing surveillance and targeted prevention and care.

## INTRODUCTION

To effectively guide patients, healthcare providers, and policymakers, it is essential to understand not only the prevalence but also the natural history and progression of long COVID symptoms following SARS-CoV-2 infection ^1^. Studies to date have provided valuable insights on understanding the natural history of long COVID symptoms ^2–13^. A prospective cohort study in Germany using healthcare system data found that fatigue, dyspnea, and reduced exercise capacity were among the most common persistent symptoms up to one year after acute SARS-CoV-2 infection ^3^. Early studies focused on electronic health records (EHR) data from individuals who accessed healthcare facilities—capturing only those who sought care—which may not reflect the full spectrum of COVID-19 severity or post-acute outcomes among people who never presented for care, thus limiting generalizability. Many studies have examined only specific dimensions of long COVID symptoms documented in EHR, which are often limited to encounter notes—resulting in a high risk of symptom omission, such as symptoms that may affect quality of life like difficulty walking. In addition, these studies often lack systematic collection and prospective follow-up, or report high attrition rates ^14^. To address these gaps, prospectively and systematically collected patient-reported outcome data across a wide range of symptom domains from community-based populations with longitudinal follow-up are essential for describing symptoms following SARS-CoV-2 infection.

Many studies now include SARS-CoV-2–uninfected comparators and adjust for key covariates, yet analytic choices often undermine causal interpretation because time zero and follow-up are misaligned between groups. When infection is contrasted with an uninfected state anchored on enrollment, calendar date, or an arbitrary survey, the resulting immortal-time and selection biases can inflate or attenuate associations—especially for symptoms with high background prevalence (e.g., fatigue, headache, cognitive complaints). Prior work has shown that adding appropriate controls (and matching on vaccination status or variant period) improves validity ^15–17^, but without explicit alignment of eligibility, index date, and outcome windows, residual bias remains ^14^. Target trial emulation (TTE) directly addresses these issues by specifying the hypothetical trial (eligibility, exposure strategies, aligned time zero, follow-up, and censoring) and then analyzing observational data accordingly ^18,19^; however, TTE has been rarely applied in studies of long COVID symptom natural history. Adopting TTE would help isolate infection effects from background symptom noise and yield more robust, policy-relevant estimates.

In this paper, we aimed to address the limitations of relying solely on EHR data and the lack of an appropriate comparison group by leveraging the CHASING COVID Cohort study, which systematically collected prospective patient-reported symptoms at multiple time points in a community-based cohort. Specifically, we characterized the natural history of long COVID symptoms and symptom clusters. Using a target trial emulation framework, we compared symptom burdens at 8 and 12 months post-acute infection between SARS-CoV-2 infected individuals and contemporaneous SARS-CoV-2 uninfected individuals ^18–20^.

## METHODS

### Participants

We recruited a geographically and socio-demographically diverse U.S. adult cohort using internet-based strategies in March 2020, with longitudinal follow-up, online surveys, and at-home dried blood spot (DBS) specimen collection ^21^. Eligibility criteria included residence in the U.S. or its territories (District of Columbia, Puerto Rico, and Guam), age ≥18 years, a valid email address for follow-up, and early study engagement (e.g., baseline specimen submission or completion of multiple recruitment visits). Detailed study design, recruitment procedures, and serology-based incidence findings have been published previously ^22–24^. Participants self-reported demographics at enrollment and approximately quarterly data on health-related characteristics—such as health insurance status, access to care, and COVID-19 vaccination status. The cohort underwent serologic testing during four periods using at-home DBS specimen kits sent and returned via the U.S. Postal Service. To detect infection-induced seroconversion, all DBS samples were tested for total antibodies to the SARS-CoV-2 nucleocapsid protein using the Bio-Rad Platelia assay for IgA, IgM, and IgG, with manufacturer-reported sensitivity of 98.0% and specificity of 99.3%, respectively ^25–27^.

### SARS-CoV-2 Infection Ascertainment, Date Assignment, and Covariate Definitions

We ascertained SARS-CoV-2 infection status using three prespecified sources of evidence: (1) self-reported positive viral tests (PCR or rapid antigen), whether provider-administered or home-based; (2) serologic evidence of prior infection from study-collected DBS specimens (total anti-nucleocapsid antibodies: IgA/IgM/IgG); and (3) the Council of State and Territorial Epidemiologists (CSTE) “probable case” definition when laboratory confirmation was unavailable ^28^. When multiple sources were present, serology was prioritized because it was systematically collected and tested within the study; otherwise, self-reported viral tests were used; if neither was available, CSTE criteria were applied. Infection dates were assigned using the most reliable information available. For self-reported positives, we used the exact date when provided, the midpoint of the reported month when only month was known, or—if neither was available—the midpoint between the report date and the previous follow-up (reflecting the survey question about positives since the last assessment). For CSTE-probable infections (used primarily during the early Omicron surge, Dec 6, 2021–Jan 11, 2022), participants reported compatible symptoms and recent epidemiologic linkage; the infection date was set to the midpoint between the assessment date and 14 days prior. Serology was fielded at four intervals (Apr–Sep 2020; Nov 2020–Mar 2021; Mar–Jun 2022; Jul–Oct 2024). Serology-identified infections lacking a corroborating test or CSTE evidence were excluded if the infection window could not be bounded within 90 days. Details are provided in Supplementary Files **Appendix 1**.

Participant characteristics included time-fixed and time-updated covariates. Time-fixed covariates (collected Mar–Jul 2020) were age (modeled with a B-spline, degree of freedom=4), self-identified gender (male, female, non-binary), race/ethnicity (Hispanic; non-Hispanic White, Black, Asian, American Indian or Pacific Islander), education, and household income (BRFSS categories ^29^). Baseline comorbidities were self-reported ever diagnoses of conditions associated with severe COVID-19 (current asthma; cancer; chronic kidney disease; chronic lung disease including COPD/emphysema/chronic bronchitis; type 2 diabetes; hypertension; heart disease; HIV; immunosuppression; and mental health conditions such as depression, PTSD, or anxiety), coded present/absent. Time-updated covariates, assessed approximately every three months, included health insurance, access to a primary care clinician (Yes/No/Don’t know), vaccination status (never; partial; completed primary series; primary plus any additional dose), and calendar month (modeled with a natural spline) to capture secular trends. Full specifications are outlined in Supplementary Files **Appendix 2**.

### Overall Study Design: Target Trial Emulation Using Sequential Trials

We used a target trial emulation framework to evaluate the effect of SARS-CoV-2 infection versus no infection on the incidence of any long COVID symptom, as well as symptoms within each of three clusters—autonomic dysfunction, exercise intolerance, and neurological symptoms—during 4-8 and 9-12 months post-infection among adult participants enrolled in the CHASING COVID Cohort in the United States. Specifications of the target trials are outlined in **Table 1**. Eligibility criteria were assessed, and exposure arms were assigned monthly from January 2021 through December 2022, resulting in 24 sequential monthly trials (**Figure 1A**). Each emulation was conducted separately for overall long COVID symptom and for each of three symptom clusters, across two follow-up durations (4–8 and 9–12 months post–time zero), resulting in eight distinct emulated trial analyses.

**Table 1.** Specification and emulation of a target trial of the SARS-CoV-2 infection and 6- and 12-month incidence of incident long COVID, autonomic dysfunction symptom, exercise intolerance symptom, and neurological symptoms among the CHASING COVID Cohort study adult participants in the U.S., January 2021-December 2022

| Protocol | Target Trial Specification | Target Trial Emulation: Cohort design |
| --- | --- | --- |
| Eligibility Criteria | **<u>Inclusion criteria:</u>**<br>•Adults aged $\geq 18$ years or older.<br>•Completion of at least one follow-up symptom survey within the four months prior to baseline.<br>**<u>Exclusion criteria:</u>**<br>•Any documented SARS-CoV-2 infection before study enrollment.<br>•First reported SARS-CoV-2 infection which the infection date cannot be reliably estimated: imputation window exceeds 90 days<br>•Classified as a CSTE-defined case for their first SARS-CoV-2 infection | Same as the target trial |
| Exposure | Participants will be assigned to:<br>1) SARS-CoV-2 infection (first infection)<br>2) No SARS-CoV-2 infection (never infected) | Same as the target trial |
| Exposure Assignment | 1) Individuals are randomly assigned to an exposure arm at baseline<br>2) Study is not blinded to investigators or enrollees | To emulate randomization via IPTW by confounders: age, gender, race/ethnicity, education, income, healthcare insurance status, access to a primary care doctor, individual comorbidity that is associate with severe COVID-19 diseases, COVID-19 vaccination receipt status, and calendar month |
| Outcomes | incidence of long COVID and 3 long COVID symptom clusters: autonomic dysfunction, exercise intolerance, and neurological symptoms, evaluated separately for 4-8 and 9-12 months post index date | Same as the target trial |
| Follow-up | In each nested trial, follow-up begins at index date of assignment and continues until earliest of<br>1) the outcome of interest<br>2) missed follow-up assessment<br>3) reinfection of SARS-CoV-2 among infected group; first infection of SARS-CoV-2 among uninfected group<br>4) administrative end of 8 or 12 months follow-up after time zero | Same as the target trial |
| Causal Contrasts | Per-protocol (PP) | Observational analogue of PP effect |
| Statistical Analysis | We will apply inverse probability censoring weights (IPCW) of pre- and post-baseline prognostic factors associated with loss to follow-up and deviation of exposure assignment: age, gender, race/ethnicity, education, income, insurance status, individual comorbidity that is associate with severe COVID-19 diseases, vaccination status, and calendar month (the time-varying covariates | Same as for the target trial with adjustment for IPTW of factors in the observational analogue of exposure assignment |
|  | will be updated to the one closest to censoring) |  |

**Figure 1.**
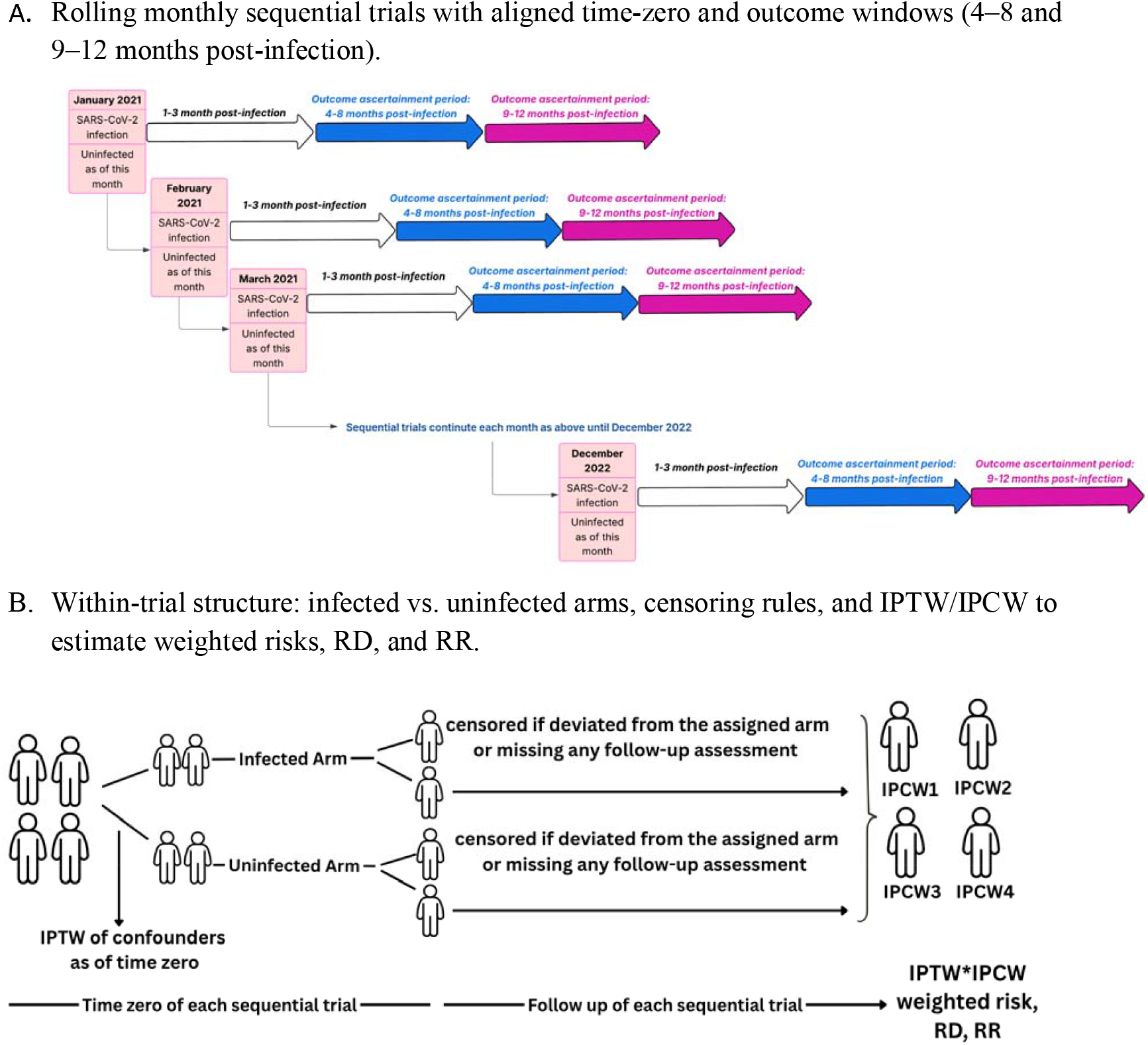
Sequential Target Trial Emulation and Weighting Strategy for Long COVID Outcomes. Panel A depicts a sequence of monthly “sequential-trials” from January 2021 through December 2022. Each month serves as an aligned time-zero at which participants are classified as infected or uninfected “as of this month.” For each monthly trial, outcomes are ascertained during two post-infection windows: 4–8 months (blue arrows) and 9– 12 months (magenta arrows). The process rolls forward each month to cover the full study period. Panel B shows the within-trial design. Two arms start at the same time-zero: an infected arm and an uninfected arm. Participants are censored if they deviate from their assigned arm or miss any scheduled follow-up assessment. Inverse probability of treatment weights (IPTW) adjust for baseline confounders at time-zero, and inverse probability of censoring weights (IPCW) adjust for selection due to censoring. The product of IPTW and IPCW is used to estimate weighted risks and to compute risk difference (RD) and risk ratio (RR) for outcomes.

#### Eligibility Criteria

For each monthly emulation, participants were potentially eligible if they were aged 18 years or older and had completed at least one follow-up assessment survey within the preceding four months. Participants were excluded if they had a documented SARS-CoV-2 infection prior to study enrollment, if the date of their first reported SARS-CoV-2 infection could not be reliably estimated, defined as requiring an imputation window exceeding 90 days or if they were classified as a CSTE-defined case for their first SARS-CoV-2 infection (**Table 1**).

#### Exposure: Ascertainment of SARS-CoV-2 infection versus no infection and assignment of index date

Between January 1^st^, 2021, and December 31^st^, 2022, individuals were classified each month as exposed if they experienced their first SARS-CoV-2 infection during that month, defined by self-reported laboratory-confirmed PCR or self-reported positive antigen test results, or a positive serology sample collected by the study. For exposed individuals, the date of first documented infection was assigned as the index date. In the same month, participants with no evidence of prior SARS-CoV-2 infection by the end of the month were identified as unexposed; for these individuals, the last day of the corresponding month was assigned as the index date.

#### Outcome

During each round of follow-up assessments conducted approximately every three months between November 2020 and December 2023, participants reported current symptoms using a standardized multiple-choice checklist (**Appendix 3**). Outcomes were evaluated separately for two follow-up intervals: 4–8 months and 9–12 months post time zero. The primary outcome was the incidence of long COVID, defined as the new onset of any long COVID symptom during the outcome measurement period that were not reported at the pre-time-zero assessment. Long COVID symptoms were selected based on the NIH RECOVER initiative’s data-driven framework, which identified core symptoms essential for characterizing long COVID (**Supplementary Table 1**) ^30^. Symptoms analyzed in this study aligned with the RECOVER definition and were selected based on their availability within the CHASING COVID Cohort study, including fatigue, trouble concentrating, brain fog, post-exertional malaise, dizziness, gastrointestinal issues, erratic heartbeat, loss or altered sense of smell, and loss or altered sense of taste.

We then defined three symptom clusters based on the NIH RECOVER sub-trials: autonomic dysfunction, exercise intolerance, and neurological symptom ^31,32^. The specific symptoms included in each cluster were outlined in **Supplementary Table 1**. The incidence definition was consistently applied to each symptom cluster individually.

#### Follow-Up

All four outcomes were evaluated separately during two distinct periods after time zero: months 4-8 and months 9-12. Participants were followed from time zero until the earliest occurrence of: (1) the outcome of interest, (2) missing a follow-up assessment, (3) reinfection among infected participants or new infection among uninfected participants, or (4) administrative censoring at 8 and 12 months after time zero.

### Statistical Analysis and Confounding Adjustment

We compared the time zero characteristics across person-trials by exposure status at time zero to assess covariate balance. Standardized mean differences (SMDs) were calculated for each covariate ^33^. We applied stabilized inverse probability of treatment and censoring weights (IPTW × IPCW) to each person-trial, then recalculated group means and variances on the weighted pseudo-populations to assess the degree to which weights reduced the imbalance.

To emulate the hypothetical randomized exposure assignment, we adjusted for covariates. These included the time-fixed variables age, gender, race, ethnicity, education, income, comorbidities that are associated with severe COVID-19 disease, and the time-updated covariates healthcare insurance status, access to a primary care doctor, COVID-19 vaccination status, and calendar month. For variables collected as time-updated measures, we used the participant’s status as of each monthly time zero; for all other variables, we used baseline data collected between March and August 2020. We employed an inverse probability treatment weighting (IPTW) approach to emulate randomization of the exposure ^34,35^.

To address potential bias due to informative loss to follow-up in either exposure group and time-varying confounding of SARS-CoV-2 infection in the no infection group, we applied artificial censoring when participants in either exposure group were lost to follow-up or the no infection group became infected and then used IPCW to reduce selection bias. Pooled logistic regression models for censoring generated the probabilities used to construct the weights at each person-time. The models used the following covariates: age, gender, race/ethnicity, education, income, healthcare insurance status, access to a primary care doctor, comorbidities that are associated with severe COVID-19 diseases, COVID-19 vaccination receipt status, and calendar month. Healthcare insurance status, access to a primary care doctor, vaccination status, and calendar month were treated as time-updated covariates, with values taken from the most recent assessment as of censoring.

At each trial-person-time, the weight was calculated as the trial time zero IPTW multiplied by the cumulative product of the inverse conditional probabilities of not being censored at each time point (**Figure 1B**). We then fit a weighted pooled logistic regression model for the discrete counterfactual hazard of outcome (i.e. symptom) occurrence in months 4-8 and separately in months 9-12 after follow-up. These models included exposure status at time zero, follow-up month (modeled as both linear and quadratic terms), and interaction terms between exposure and follow-up month to allow for non-proportional effects over time. Survival probabilities were obtained as the cumulative product of one minus the estimated hazard at each time point, and risks were defined as one minus the survival probability. We estimated the cumulative incidence, risk differences, and risk ratios for each outcome and each time frame, comparing infected and uninfected individuals. We calculated 95% confidence intervals (CIs) via nonparametric bootstrapping with 500 replicates, resampling at the participant level (i.e., all person□trials for a given individual were drawn together in each replicate) ^36^. In a sensitivity analysis, we included CSTE-defined probable infections when assigning the exposure arm; per-protocol IPTW/IPCW estimates (weighted incidence, incidence difference, and incidence ratio at 4–8 and 9–12 months) are reported in Supplementary Table 4. We used SAS version 9.4 (SAS Institute, Cary, NC, USA) for data management and R version 2024.09.0+375 (R Foundation for Statistical Computing, Vienna, Austria) for data analyses.

## RESULTS

### Comparison of Characteristics of SARS-CoV-2 Infected and Uninfected Individuals

A total of 24 sequentially emulated trials were included in the analysis, comprising 1,055 exposed (infected) and 52,310 unexposed (uninfected) person-trials. **Table 2** summarizes the baseline characteristics of infected and uninfected individuals across these trials. Overall, the two groups were generally well balanced across most sociodemographic and clinical characteristics, as indicated by standardized mean differences (SMDs) below 0.2 for the majority of covariates. A large imbalance was observed for age (SMD = 0.286), with infected individuals more likely to be younger. COVID-19 vaccination status showed substantial imbalance (SMD = 0.516), with a higher proportion of uninfected individuals reporting full or partial vaccination. Modest imbalances were also observed for hypertension (SMD = 0.120), chronic lung disease (SMD = 0.076), and heart disease (SMD = 0.073). Other characteristics—including gender, race, ethnicity, education, income, insurance status, and most comorbidities—were similarly distributed across groups. All SMDs were below 0.1 in the weighted pseudo population, indicating adequate balance.

**Table 2.**
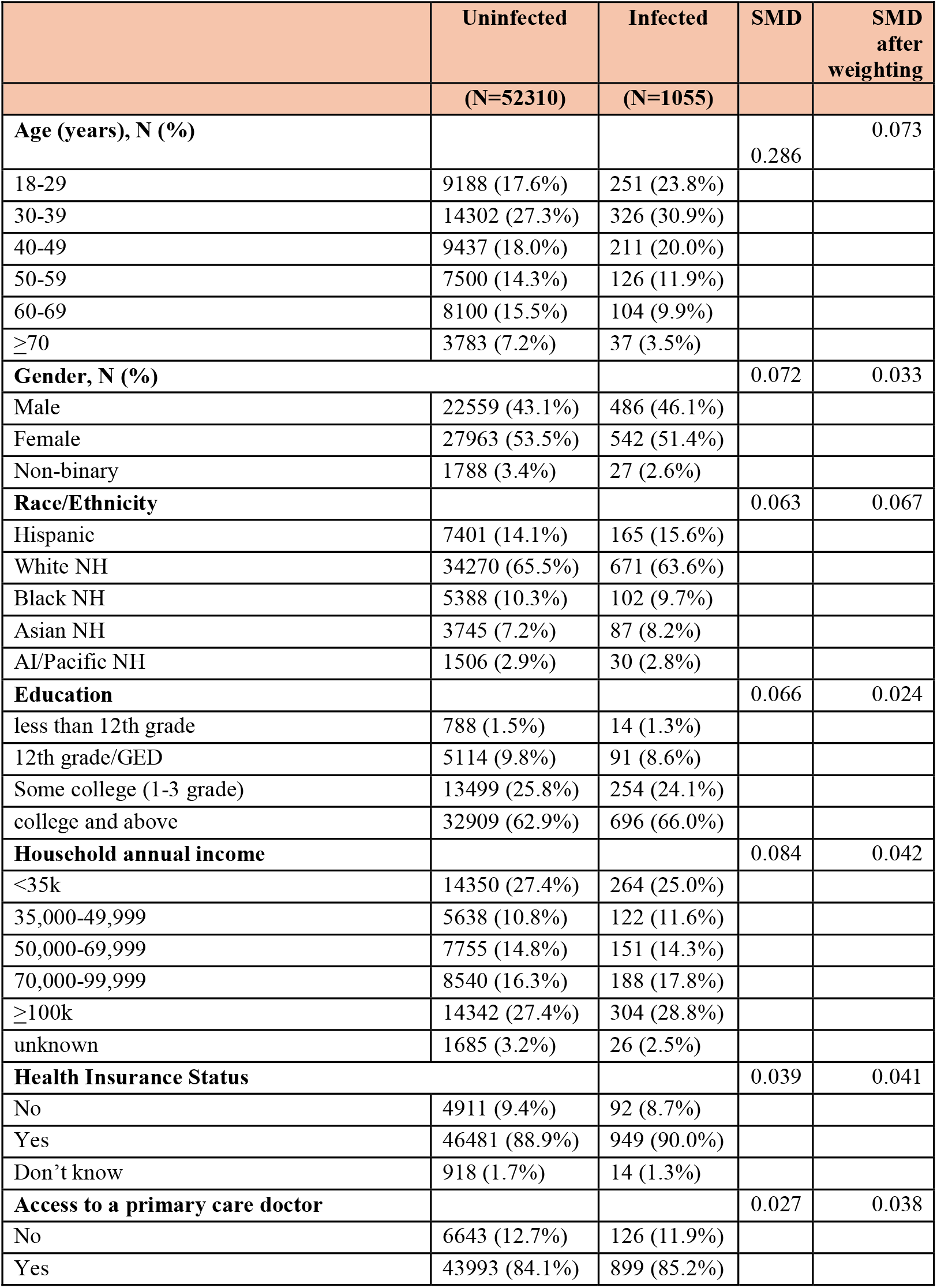
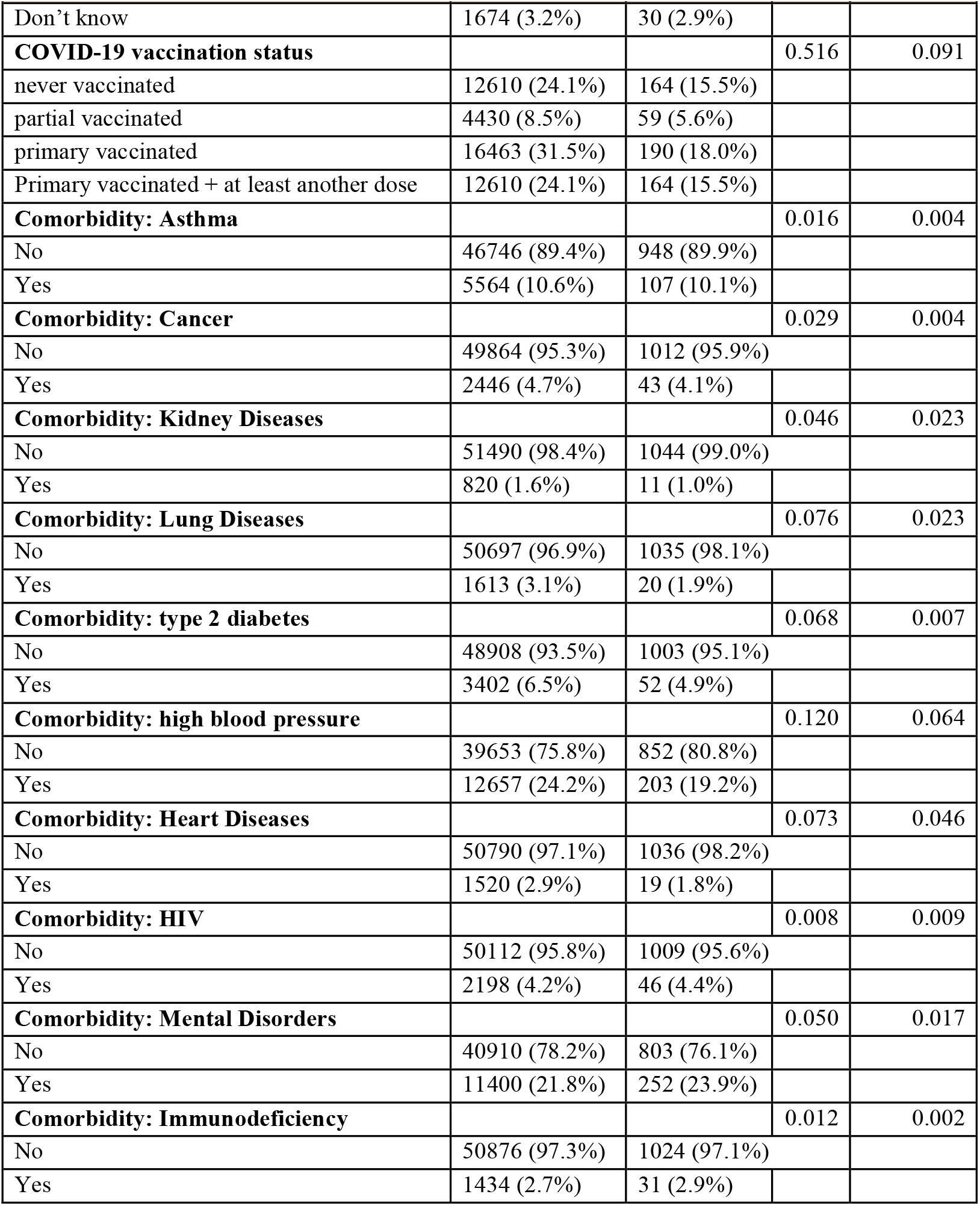
Characteristics of infected verse uninfected individuals across 24 sequential trails.

|  | Uninfected | Infected | SMD | SMD<br>after<br>weighting |
| --- | --- | --- | --- | --- |
|  | (N=52310) | (N=1055) |  |  |
| **Age (years), N (%)** |  |  | 0.286 | 0.073 |
| 18-29 | 9188 (17.6%) | 251 (23.8%) |  |  |
| 30-39 | 14302 (27.3%) | 326 (30.9%) |  |  |
| 40-49 | 9437 (18.0%) | 211 (20.0%) |  |  |
| 50-59 | 7500 (14.3%) | 126 (11.9%) |  |  |
| 60-69 | 8100 (15.5%) | 104 (9.9%) |  |  |
| ≥70 | 3783 (7.2%) | 37 (3.5%) |  |  |
| **Gender, N (%)** |  |  | 0.072 | 0.033 |
| Male | 22559 (43.1%) | 486 (46.1%) |  |  |
| Female | 27963 (53.5%) | 542 (51.4%) |  |  |
| Non-binary | 1788 (3.4%) | 27 (2.6%) |  |  |
| **Race/Ethnicity** |  |  | 0.063 | 0.067 |
| Hispanic | 7401 (14.1%) | 165 (15.6%) |  |  |
| White NH | 34270 (65.5%) | 671 (63.6%) |  |  |
| Black NH | 5388 (10.3%) | 102 (9.7%) |  |  |
| Asian NH | 3745 (7.2%) | 87 (8.2%) |  |  |
| AI/Pacific NH | 1506 (2.9%) | 30 (2.8%) |  |  |
| **Education** |  |  | 0.066 | 0.024 |
| less than 12th grade | 788 (1.5%) | 14 (1.3%) |  |  |
| 12th grade/GED | 5114 (9.8%) | 91 (8.6%) |  |  |
| Some college (1-3 grade) | 13499 (25.8%) | 254 (24.1%) |  |  |
| college and above | 32909 (62.9%) | 696 (66.0%) |  |  |
| **Household annual income** |  |  | 0.084 | 0.042 |
| <35k | 14350 (27.4%) | 264 (25.0%) |  |  |
| 35,000-49,999 | 5638 (10.8%) | 122 (11.6%) |  |  |
| 50,000-69,999 | 7755 (14.8%) | 151 (14.3%) |  |  |
| 70,000-99,999 | 8540 (16.3%) | 188 (17.8%) |  |  |
| ≥100k | 14342 (27.4%) | 304 (28.8%) |  |  |
| unknown | 1685 (3.2%) | 26 (2.5%) |  |  |
| **Health Insurance Status** |  |  | 0.039 | 0.041 |
| No | 4911 (9.4%) | 92 (8.7%) |  |  |
| Yes | 46481 (88.9%) | 949 (90.0%) |  |  |
| Don't know | 918 (1.7%) | 14 (1.3%) |  |  |
| **Access to a primary care doctor** |  |  | 0.027 | 0.038 |
| No | 6643 (12.7%) | 126 (11.9%) |  |  |
| Yes | 43993 (84.1%) | 899 (85.2%) |  |  |
|  | (N=52310) | (N=1055) |  |  |
| Don't know | 1674 (3.2%) | 30 (2.9%) |  |  |
| **COVID-19 vaccination status** |  |  | 0.516 | 0.091 |
| never vaccinated | 12610 (24.1%) | 164 (15.5%) |  |  |
| partial vaccinated | 4430 (8.5%) | 59 (5.6%) |  |  |
| primary vaccinated | 16463 (31.5%) | 190 (18.0%) |  |  |
| Primary vaccinated + at least another dose | 12610 (24.1%) | 164 (15.5%) |  |  |
| **Comorbidity: Asthma** |  |  | 0.016 | 0.004 |
| No | 46746 (89.4%) | 948 (89.9%) |  |  |
| Yes | 5564 (10.6%) | 107 (10.1%) |  |  |
| **Comorbidity: Cancer** |  |  | 0.029 | 0.004 |
| No | 49864 (95.3%) | 1012 (95.9%) |  |  |
| Yes | 2446 (4.7%) | 43 (4.1%) |  |  |
| **Comorbidity: Kidney Diseases** |  |  | 0.046 | 0.023 |
| No | 51490 (98.4%) | 1044 (99.0%) |  |  |
| Yes | 820 (1.6%) | 11 (1.0%) |  |  |
| **Comorbidity: Lung Diseases** |  |  | 0.076 | 0.023 |
| No | 50697 (96.9%) | 1035 (98.1%) |  |  |
| Yes | 1613 (3.1%) | 20 (1.9%) |  |  |
| **Comorbidity: type 2 diabetes** |  |  | 0.068 | 0.007 |
| No | 48908 (93.5%) | 1003 (95.1%) |  |  |
| Yes | 3402 (6.5%) | 52 (4.9%) |  |  |
| **Comorbidity: high blood pressure** |  |  | 0.120 | 0.064 |
| No | 39653 (75.8%) | 852 (80.8%) |  |  |
| Yes | 12657 (24.2%) | 203 (19.2%) |  |  |
| **Comorbidity: Heart Diseases** |  |  | 0.073 | 0.046 |
| No | 50790 (97.1%) | 1036 (98.2%) |  |  |
| Yes | 1520 (2.9%) | 19 (1.8%) |  |  |
| **Comorbidity: HIV** |  |  | 0.008 | 0.009 |
| No | 50112 (95.8%) | 1009 (95.6%) |  |  |
| Yes | 2198 (4.2%) | 46 (4.4%) |  |  |
| **Comorbidity: Mental Disorders** |  |  | 0.050 | 0.017 |
| No | 40910 (78.2%) | 803 (76.1%) |  |  |
| Yes | 11400 (21.8%) | 252 (23.9%) |  |  |
| **Comorbidity: Immunodeficiency** |  |  | 0.012 | 0.002 |
| No | 50876 (97.3%) | 1024 (97.1%) |  |  |
| Yes | 1434 (2.7%) | 31 (2.9%) |  |  |

### Incidence of Long COVID Symptoms

**Table 3a** presents the IPTW- and IPCW-weighted incidence, risk differences, and risk ratios for long COVID symptoms comparing SARS-CoV-2 infected and uninfected individuals across two follow-up intervals. At 4–8 months post time zero, the adjusted risk of experiencing any long COVID symptom was 22.6% (95% CI: 20.5%, 24.8%) among infected individuals, compared to % (95% CI: 11.1%, 11.5%) among uninfected individuals. This yielded an adjusted risk difference (aRD) of 11.3% (95% CI: 9.2%, 13.5%) and an adjusted risk ratio (aRR) of 2.005 (95% CI: 1.809, 2.197). At 9–12 months, the adjusted risk was 19.2% (95% CI: 17.0%, 21.3%) among infected participants and 12.4% (95% CI: 12.2%, 12.7%) among uninfected participants. The corresponding aRD was 6.7% (95% CI: 4.6%, 8.9%), and the aRR was 1.542 (95% CI: 1.366, 1.720).

**Table 3.** Analog of the per-protocol IPTW and IPCW weighted incidence, incidence difference, and incidence ratio (95% confidence intervals) for 4-8 and 9-12-month of incident long COVID symptoms, The CHASING COVID Cohort, January 2021 – December 2023.

| Follow up time | Exposure Arm | IPTW and IPCW-adjusted risk (95% CI) |  | Risk difference (95% CI) |  | Risk ratio (95% CI) |  |
| --- | --- | --- | --- | --- | --- | --- | --- |
| 4-8 months | Participants with SARS-CoV-2 infection | 0.226 | 0.205, 0.248 | 0.113 | 0.092, 0.135 | 2.005 | 1.809, 2.197 |
|  | Participants without SARS-CoV-2 infection | 0.113 | 0.111, 0.115 | ref | ref | ref | ref |
| 9-12 months | Participants with SARS-CoV-2 infection | 0.192 | 0.170, 0.213 | 0.067 | 0.046, 0.089 | 1.542 | 1.366, 1.720 |
|  | Participants without SARS-CoV-2 infection | 0.124 | 0.122, 0.127 | ref | ref | ref | ref |

b) Long COVID with autonomic dysfunction symptoms
| Follow up time | Exposure Arm | IPTW and IPCW-adjusted risk (95% CI) |  | Risk difference (95% CI) |  | Risk ratio (95% CI) |  |
| --- | --- | --- | --- | --- | --- | --- | --- |
| 4-8 months | Participants with SARS-CoV-2 infection | 0.104 | 0.088, 0.118 | 0.043 | 0.031, 0.057 | 1.700 | 1.501, 1.875 |
|  | Participants without SARS-CoV-2 infection | 0.061 | 0.059, 0.063 | ref | ref | ref | ref |
| 9-12 months | Participants with SARS-CoV-2 infection | 0.098 | 0.081, 0.113 | 0.032 | 0.019, 0.048 | 1.479 | 1.267, 1.665 |
|  | Participants without SARS-CoV-2 infection | 0.066 | 0.064, 0.068 | ref | ref | ref | ref |

c) Long COVID with exercise intolerance symptoms
| Follow up time | Exposure Arm | IPTW and IPCW-adjusted risk (95% CI) |  | Risk difference (95% CI) |  | Risk ratio (95% CI) |  |
| --- | --- | --- | --- | --- | --- | --- | --- |
| 4-8 months | Participants with SARS-CoV-2 infection | 0.137 | 0.121, 0.153 | 0.072 | 0.054, 0.087 | 2.100 | 1.921, 2.287 |
|  | Participants without SARS-CoV-2 infection | 0.065 | 0.063, 0.067 | ref | ref | ref | ref |
| 9-12 months | Participants with SARS-CoV-2 infection | 0.119 | 0.102, 0.137 | 0.046 | 0.030, 0.062 | 1.622 | 1.418, 1.805 |
|  | Participants without SARS-CoV-2 infection | 0.074 | 0.072, 0.076 | ref | ref | ref | ref |

d) Long COVID with neurologic symptoms
| Follow up time | Exposure Arm | IPTW and IPCW-adjusted risk (95% CI) |  | Risk difference (95% CI) |  | Risk ratio (95% CI) |  |
| --- | --- | --- | --- | --- | --- | --- | --- |
| 4-8 months | Participants with SARS-CoV-2 infection | 0.142 | 0.126, 0.158 | 0.067 | 0.055, 0.083 | 1.899 | 1.721, 2.056 |
|  | Participants without SARS-CoV-2 infection | 0.075 | 0.073, 0.077 | ref | ref | ref | ref |
| 9-12 months | Participants with SARS-CoV-2 infection | 0.121 | 0.104, 0.139 | 0.039 | 0.022, 0.053 | 1.481 | 1.298, 1.659 |
|  | Participants without SARS-CoV-2 infection | 0.082 | 0.080, 0.084 | ref | ref | ref | ref |

### Incidence of Symptom Clusters

Across all three symptom clusters—autonomic dysfunction, exercise intolerance, and neurological symptoms—infected individuals consistently showed higher adjusted risks compared to uninfected individuals (**Table 3b-3d**). For autonomic dysfunction symptoms (**Table 3b**), the risk at 4–8 months was 10.4% (95% CI: 8.8%, 11.8%) among infected individuals versus 6.1% (95% CI: 5.9%, 6.3%) among uninfected individuals, yielding an aRD of 4.3% (95% CI: 3.1%, 5.7%) and an aRR of 1.700 (95% CI: 1.501, 1.875). At 9–12 months, the risk among infected individuals was 9.8% (95% CI: 8.1%, 11.3%), compared to 6.6% (95% CI: 6.4%, 6.8%) among uninfected individuals (aRD: 3.2% [95% CI: 1.9%, 4.8%]; aRR: 1.479 [95% CI: 1.267, 1.665]).

For exercise intolerance symptoms (**Table 3c**), the 4–8 month risk was 13.7% (95% CI: 12.1%, 15.3%) among infected individuals and 6.5% (95% CI: 6.3%, 6.7%) among uninfected individuals. The associated aRD was 7.2% (95% CI: 5.4%, 8.7%) and the aRR was 2.100 (95% CI: 1.921, 2.287). At 9–12 months, the risk was 11.9% (95% CI: 10.2%, 13.7%) in the infected group versus 7.4% (95% CI: 7.2%, 7.6%) in the uninfected group (aRD: 4.6% [95% CI: 3.0%, 6.2%]; aRR: 1.622 [95% CI: 1.418, 1.805]).

For neurological symptoms (**Table 3d**), the adjusted risk at 4–8 months was 14.2% (95% CI: 12.6%, 15.8%) among infected individuals compared to 7.5% (95% CI: 7.3%, 7.7%) among uninfected individuals. This resulted in an aRD of 6.7% (95% CI: 5.5%, 8.3%) and an aRR of 1.899 (95% CI: 1.721, 2.056). At 9–12 months, the risk was 12.1% (95% CI: 10.4%, 13.9%) among infected individuals and 8.2% (95% CI: 8.0%, 8.4%) among uninfected individuals, corresponding to an aRD of 3.9% (95% CI: 2.2%, 5.3%) and an aRR of 1.481 (95% CI: 1.298, 1.659).

## DISCUSSION

Our prospective community-based study evaluated the natural history of symptoms following SARS-CoV-2 infection, up to 12 months post-infection, relative to individuals who were contemporaneously SARS-CoV-2 uninfected. Our findings aligned with prior research highlighting the prolonged nature of post-COVID symptoms and an observed decline over time. A retrospective cohort study utilizing EHR data described a gradual decline in symptom burden among COVID-19 survivors, with the highest prevalence during the acute phase and a slow recovery over subsequent months, within the first year post-infection ^2^. Similarly, in a prospective e-cohort study of COVID-19 survivors, a substantial proportion of individuals continued to report symptoms more than six months after infection, with some lasting for up to a year^6^. Together, these studies support the persistence of post-COVID symptoms and align with the trends observed in our cohort. Importantly, by including a contemporaneous SARS-CoV-2 uninfected control group, our study enhances prior findings by isolating the impact of SARS-CoV-2 infection from background symptom prevalence in the general population—offering a more precise assessment of infection-attributable symptom burden. In addition, our results suggest that the risk of new-onset symptoms—while still elevated at 12 months—may decline between the 4–8 and 9–12 month follow-up periods after infection. This trend added to the understanding of symptom trajectories and reinforces the importance of characterizing symptom evolution over time in natural history studies.

Our study reinforced the substantial burden of neurological, autonomic dysfunction and exercise-intolerance symptoms among individuals with SARS-CoV-2 infection compared to those who were contemporaneously SARS-CoV-2 negative at both follow-up intervals. Seeßle et al. and Rass et al. reported neurological deficits, including cognitive dysfunction and fatigue, lasting up to one year post-infection ^3,5^. Dennis et al. also identified long-term multi-organ impairment, particularly affecting autonomic function, in long COVID patients ^7^. Furthermore, Wu et al. described prolonged respiratory complications following hospitalization, which aligns with our findings on exercise intolerance ^13^. While absolute risks declined slightly over time, elevated risks for exercise intolerance, autonomic dysfunction, and neurological symptoms remained elevated through 12 months post-infection.

Our study has limitations. First, we could not distinguish mild versus severe acute COVID-19, limiting assessment of how acute severity shapes long COVID trajectories. Second, the symptom inventory was predefined and omitted some relevant features (e.g., chest pain; limb weakness; skin color changes, dry mouth; confusion/forgetfulness), potentially underestimating domain-specific incidence. Third, symptoms were self-reported, introducing recall/reporting bias; however, self-report is necessary for symptoms (e.g., fatigue) that are not readily elicited clinically. Fourth, despite inverse probability weighting, residual confounding from measured and unmeasured factors (e.g., baseline inflammation, genetic susceptibility, environmental exposures) remains possible. Fifth, infection timing is imperfect: although we excluded cases with >90-day imputation windows and those classified solely as probable by CSTE criteria, some misclassification likely persists, affecting estimates of onset and duration. Sixth, in the absence of a standardized long COVID case definition, we used an expert-informed, symptom-based definition; while broadly aligned with NIH RECOVER, differences in instruments and timing preclude assessing sensitivity/specificity and limit cross-study comparability. Finally, comorbidities were self-reported and not validated against records; our immunodeficiency measure may miss therapy-induced immunosuppression, and conditions were reported as “ever”, preventing differentiation of active versus resolved disease and likely attenuating associations.

In summary, we found a high proportion of new onset long COVID symptoms—especially neurological, autonomic dysfunction, and exercise-intolerance features—persisting up to 12 months post-infection, compared with contemporaneous SARS-CoV-2–negative individuals. Using a national community-based cohort with contemporaneous controls and symptom clustering, we provided a more nuanced view of post-acute symptom natural history than EHR-based studies. These findings reinforce the heterogeneity and persistence of long COVID and support targeted interventions—including rehabilitation, autonomic dysfunction management, cognitive therapy, and exercise-based programs—to mitigate long-term symptoms and improve outcomes ^37–41^.

## PATIENT CONSENT STATEMENT

Informed consent forms were completed in a web browser on participants’ computer or mobile device at baseline, each round of serological testing, and at periodic follow-up assessments. The study was approved by the Institutional Review Boards of the City University Of New York (CUNY) Graduate School of Public Health and Health Policy (New York, NY, U.S.) (protocol 2020-0256).

## ACKNOWLEDGEMENTS

The authors wish to thank the participants of the CHASING COVID Cohort Study. We are grateful to you for your contributions to the advancement of science around the SARS-CoV-2 pandemic.

## AUTHOR CONTRIBUTORS

YS: Conceptualization; Study design; Data curation; Formal analysis; Investigation; Visualization; Writing—original draft; Writing—review & editing. DN: Conceptualization; Supervision; Resources; Funding acquisition; Project administration; Writing—review & editing. ZS: Methodology; Interpretation; Writing—review & editing. MR: Methodology; Interpretation; Writing—review & editing. KG: Methodology; Interpretation; Writing—review & editing.

## DATA AVAILABILITY

The data underlying this study contain potentially identifiable health information and are not publicly available due to ethical and legal restrictions. De-identified data may be shared on reasonable request and subject to institutional approvals and a Data Use Agreement. Requests will be reviewed case-by-case by the study investigators and the CUNY ISPH data governance/IRB. Please contact the corresponding author.

