## supplementary files for "Natural History of Self-reported Symptoms Following SARS-CoV-2 Infection: A Target Trial Emulation in a Prospective Community-Based Cohort"

### Appendix Table 1: STROBE (Strengthening the Reporting of Observational studies with Epidemiology) Checklist of items that should be included in reports of cohort studies.

|  | Item | Recommendation | Page |
| --- | --- | --- | --- |
| **Title and abstract** | 1 | (a) Indicate the study’s design with a commonly used term in the title or the abstract |  |
|  |  | (b) Provide in the abstract an informative and balanced summary of what was done and what was found | 1-2 |
| Introduction | | | |
| Background/rationale | 2 | Explain the scientific background and rationale for the investigation being reported | 3 |
| Objectives | 3 | State specific objectives, including any prespecified hypotheses | 3 |
| Methods | | | |
| Study design | 4 | Present key elements of study design early in the paper | 4 |
| Setting | 5 | Describe the setting, locations, and relevant dates, including periods of recruitment, exposure, follow-up, and data collection | 4 |
| Participants | 6 | (a) Give the eligibility criteria, and the sources and methods of selection of participants. Describe methods of follow-up |  |
|  |  | (b) For matched studies, give matching criteria and number of exposed and unexposed | 5, Table 1 |
| Variables | 7 | Clearly define all outcomes, exposures, predictors, potential confounders, and effect modifiers. Give diagnostic criteria, if applicable | 5-6 |
| Data sources/ measurement | 8 | For each variable of interest, give sources of data and details of methods of assessment (measurement). Describe comparability of assessment methods if there is more than one group | 5-6 |
| Bias | 9 | Describe any efforts to address potential sources of bias | 4-6 |
| Study size | 10 | Explain how the study size was arrived at | Supplementary Figure 3 |
| Quantitative variables | 11 | Explain how quantitative variables were handled in the analyses. If applicable, describe which groupings were chosen and why | 3-5, Appendix 1, 2 |
| Statistical methods | 12 | (a) Describe all statistical methods, including those used to control for confounding |  |
|  |  | (b) Describe any methods used to examine subgroups and interactions | 6-7 |
|  |  | (c) Explain how missing data were addressed |  |
|  |  | (d) If applicable, explain how loss to follow-up was addressed |  |
|  |  | (e) Describe any sensitivity analyses |  |
| Results | | |  |
| Participants | 13 | (a) Report numbers of individuals at each stage of study—eg numbers potentially eligible, examined for eligibility, confirmed eligible, included in the study, completing follow-up, and analyzed | 7, Supplementary Figure 3 |
|  |  | (b) Give reasons for non-participation at each stage |  |
|  |  | (c) Consider use of a flow diagram |  |
| Descriptive data | 14 | (a) Give characteristics of study participants (eg demographic, clinical, social) and information on exposures and potential confounders | 6-7, Table 2 |
|  |  | (b) Indicate number of participants with missing data for each variable of interest |  |
|  |  | (c) Summarize follow-up time (eg, average and total amount) |  |
| Outcome data | 15 | Report numbers of outcome events or summary measures over time | 7-8, Table 3 |
| Main results | 16 | (a) Give unadjusted estimates and, if applicable, confounder-adjusted estimates and their precision (eg, 95% confidence interval). Make clear which confounders were adjusted for and why they were included  (b) Report category boundaries when continuous variables were categorized  (c) If relevant, consider translating estimates of relative risk into absolute risk for a meaningful time period | 7-8, Table 3 |
| Other analyses | 17 | Report other analyses done—eg analyses of subgroups and interactions, and sensitivity analyses | Supplementary Table 4 |
| **Discussion** |  |  |  |
| Key results | 18 | Summarise key results with reference to study objectives | 8 |
| Limitations | 19 | Discuss limitations of the study, taking into account sources of potential bias or imprecision. Discuss both direction and magnitude of any potential bias | 9 |
| Interpretation | 20 | Give a cautious overall interpretation of results considering objectives, limitations, multiplicity of analyses, results from similar studies, and other relevant evidence | 8-9 |
| Generalisability | 21 | Discuss the generalisability (external validity) of the study results | 9 |
| **Other information** |  |  |  |
| Funding | 22 | Give the source of funding and the role of the funders for the present study and, if applicable, for the original study on which the present article is based | 10 |

### Appendix 1: SARS-CoV-2 Infection and Dates

Infection status was classified based on one or more of the following criteria: (1) a self-reported positive result from a polymerase chain reaction (PCR) or rapid antigen test, regardless of whether it was administered by a healthcare provider or completed at home; (2) serologic evidence of prior infection, indicated by the detection of total antibodies (IgA, IgM, or IgG) against the SARS-CoV-2 nucleocapsid protein from dried blood spot (DBS) specimens collected through the study; or (3) classification as a probable case according to the Council of State and Territorial Epidemiologists (CSTE) definition. In cases where multiple sources of evidence were available, serologic evidence was prioritized for determining infection status, as it was systematically collected and tested through the study. If serology was unavailable, we relied on self-reported viral test results for infection classification. When neither of these were available, participants were classified as infected based on the CSTE probable case definition. Infection dates were assigned based on the most reliable evidence available, following the methods outlined below. Infection dates were determined by a self-reported exact date or were imputed based on the earliest and latest plausible dates of infection—creating an infection window bounded by the date of survey collection, reported symptoms and epidemiologic linkage, or serologic sample collection, depending on the infection classification method.

(1) Infection Identified by Positive PCR or Antigen Tests: SARS-CoV-2 infection was estimated through self-reported positive viral PCR or antigen test, either conducted by a healthcare provider or using a home-based rapid test. When participants provided an exact date, that date was used; if only the month without a specific day was reported, the midpoint of that month was assigned as the infection date. When the exact infection date or the month of infection is unavailable, infection date was imputed as the midpoint between the date they reported the infection and the previous follow-up assessment, reflecting the question about positive viral tests since the last survey.

(2) Infections Identified by CSTE Criteria: Given the limited testing capacity during the initial surge of Omicron infections in the U.S. ^1–4^, we identified probable infections occurring between December 6, 2021 and January 11, 2022 using the Council of State and Territorial Epidemiologists (CSTE) probable case definition ^5^. Participants reported: 1) experiencing either at least one of the following symptoms — cough, shortness of breath, loss or altered sense of smell, loss or altered sense of taste, or chest pain; or at least two of the following symptoms — fever, chills, myalgia, headache, sore throat, nausea/vomiting, congestion/runny nose; and 2) having epidemiologic linkage in the prior 14 days, including close contact with a confirmed or probable SARS-CoV-2 case or being a healthcare worker. Since the epidemiologic linkage was assessed as two weeks prior to the follow-up assessment, the infection date was assigned as the midpoint between the assessment date and 14 days prior.

(3) Infections Identified by Positive Serology Tests: We also identified SARS-CoV-2 infections using serologic testing results from study-collected dried blood spots (DBS) at 4 time points: April-September 2020 (Serology Period 1), November 2020-March 2021 (Serology Period 2), March-June 2022 (Serology Period 3), and July 2024-October 2024 (Serology Period 4). For serology-identified infections accompanied by a self-reported positive PCR or antigen test, or meeting CSTE criteria, infection dates were determined using the methods described. Serology-identified infections without a prior viral or serology test or CSTE evidence were excluded, as infection dates could not be reliably estimated within a 90-day interval.

### Appendix 2: Demographic and Comorbidity Characteristics Definitions

Time-fix covariates age, self-identified gender, race, ethnicity, education, and household annual income were collected between March and July 2020. Continuous age was modeled using B-spline with 4 degrees of freedom. Self-identified gender was grouped as male, female, and non-binary. Race/ethnicity was categorized as Hispanic, non-Hispanic White, non-Hispanic Black, non-Hispanic Asian, and non-Hispanic American Indian or Pacific Islander. Income was categorized based on self-reported annual household income brackets according to BRFSS categories. Participants self-reported whether they had ever been diagnosed with any of the conditions identified by the CDC as risk factors for severe COVID-19 illness, using a checklist allowing multiple selection collected at study enrollment. Conditions included current asthma, cancer, chronic kidney disease (not including kidney stones, bladder infection or incontinence), chronic lung disease (including COPD, emphysema, or chronic bronchitis), type 2 diabetes, high blood pressure, heart disease (including myocardial infarction, angina or coronary heart disease), HIV, immunosuppression, and mental health conditions (including depression, PTSD, or anxiety) ^6^. All conditions were coded as present or absent.

Time-updated covariates health insurance status, access to a primary care doctor, COVID-19 vaccination status, and comorbidities were collected over the course of follow up assessment roughly every three months. Health insurance status and access to a primary care doctor were categorized as “Yes,” “No,” or “Don’t know,” based on response options provided in the survey. COVID-19 vaccination status was coded as “Never vaccinated” if they had received no doses, “Partially vaccinated” if they had initiated but not completed a primary vaccine series, “Completed a primary vaccine series” if they had received the full initial series (e.g., 2 doses of an mRNA vaccine or 1 dose of the J&J vaccine), and “Primary Vaccine plus any additional dose” if they had received at least one booster or additional dose beyond the primary series. Calendar month, used to account for secular trends in infection risk and outcome reporting, was treated as a continuous variable modeled using a natural spline term.

Appendix 3: Symptom assessment question

Are you currently experiencing any of the following symptoms? Please select all that apply.

a) Shortness of breath

b) Difficulty walking more than 15 minutes

c) Difficulty running / exercising

d) Fatigue

e) Fatigue after an activity (e.g., doing dishes, which is sometimes called post exertional malaise)

f) Headache

g) Trouble concentrating / brain fog

h) Dizziness

i) Irritability

j) Erratic heartbeat

k) Gastro-intestinal issues

l) Low-grade fever

m) Muscle aches (myalgia)

n) Loss or altered taste

o) Loss or altered sense of smell

p) Waxing and waning of some or all of my initial symptoms

q) Difficulty sleeping

r) Something else:_____

s) I am NOT experiencing any of the symptoms above [exclusive]

### Supplementary Table 1: Symptoms aligned with RECOVER trials and symptom variable definitions.

*Reference:

| **RECOVER definition/cluster *** | **Symptoms used in the RECOVER studies** | **CHASING COVID Cohort study collected symptoms** |
| --- | --- | --- |
| **Long COVID symptoms** | Smell loss | Loss or altered sense of smell |
|  | Taste loss | Loss or altered sense of taste |
|  | Post-exertional malaise | Post-exertional malaise |
|  | Chronic cough | Not collected |
|  | Brain fog | Trouble concentrating/brain fog |
|  | Thirst | Not collected |
|  | Palpitations | Erratic Heartbeat |
|  | Chest Pain | Not collected |
|  | Fatigue | Fatigue |
|  | Sexual desire or capacity | Not collected |
|  | Dizziness | Dizziness |
|  | Gastrointestinal | Gastro-intestinal issues |
|  | Abnormal movements | Not collected |
|  | Hair loss | Not collected |
| **Exercise Intolerance** | Fatigue (being tired) | Fatigue |
|  | Post-exertional malaise (symptoms worse after even minor physical or mental effort) | Post-exertional malaise |
|  | Weakness in your arms and/or legs | Not collected |
| **Autonomic Dysfunction** | Pain in your limbs | Myalgia |
|  | Faintness (light-headedness) | Dizziness |
|  | Gastrointestinal (GI) upset | Gastrointestinal issues |
|  | Color changes of the skin | Not collected |
|  | Dry mouth | Not collected |
| **Neurological Symptoms** | Problems with thinking or concentrating (brain fog) | Trouble concentrating/brain fog |
|  | Confusion | Not collected |
|  | Forgetfulness | Not collected |
|  | Difficulty focusing | Trouble concentrating/brain fog |
|  | Difficulty sleeping | Difficulty sleeping |

Thaweethai T, Jolley SE, Karlson EW, et al. Development of a definition of postacute sequelae of SARS-CoV-2 infection. JAMA. 2023;329(22):1934-1946.

RECOVER-VITAL: A Platform Protocol for Evaluation of Interventions for Viral Persistence, Viral Reactivation, and Immune Dysregulation in Post-Acute Sequelae of SARS-CoV-2 Infection (PASC). (https://clinicaltrials.gov/study/NCT05965726?cond=Long&tab=table)

### Supplementary Table 2: Pre-infection symptom comparing infected and uninfected individuals across 24 sequential trials

|  | Infected individuals | Uninfected individuals | SMD | SMD after weighting |
| --- | --- | --- | --- | --- |
|  | **(N=52310)** | **(N=1055)** |  |  |
| **Pre-time-zero shortness of breath** |  |  | 0.043 | 0.019 |
| No | 50485 (96.5%) | 1026 (97.3%) | |  |
| Yes | 1825 (3.5%) | 29 (2.7%) |  |  |
| **Pre-time-zero Difficulty walking** |  |  | 0.081 | 0.046 |
| No | 50279 (96.1%) | 1029 (97.5%) | |  |
| Yes | 2031 (3.9%) | 26 (2.5%) |  |  |
| **Pre-time-zero Difficulty Running** |  |  | 0.119 | 0.096 |
| No | 49738 (95.1%) | 1027 (97.3%) | |  |
| Yes | 2572 (4.9%) | 28 (2.7%) |  |  |
| **Pre-time-zero Fatigue** |  |  | 0.002 | 0.006 |
| No | 46078 (88.1%) | 930 (88.2%) |  |  |
| Yes | 6232 (11.9%) | 125 (11.8%) |  |  |
| **Pre-time-zero Headache** | |  | 0.011 | 0.026 |
| No | 47672 (91.1%) | 958 (90.8%) |  |  |
| Yes | 4638 (8.9%) | 97 (9.2%) |  |  |
| **Pre-time-zero Trouble concentrating** |  |  | 0.035 | 0.044 |
| No | 48626 (93.0%) | 971 (92.0%) |  |  |
| Yes | 3684 (7.0%) | 84 (8.0%) |  |  |
| **Pre-time-zero Dizziness** |  |  | 0.011 | 0.029 |
| No | 50726 (97.0%) | 1021 (96.8%) | |  |
| Yes | 1584 (3.0%) | 34 (3.2%) |  |  |
| **Pre-time-zero Irritability** |  |  | 0.003 | 0.005 |
| No | 49101 (93.9%) | 991 (93.9%) |  |  |
| Yes | 3209 (6.1%) | 64 (6.1%) |  |  |
| **Pre-time-zero Erratic Heartbeat** |  |  | 0.013 | 0.023 |
| No | 51454 (98.4%) | 1036 (98.2%) | |  |
| Yes | 856 (1.6%) | 19 (1.8%) |  |  |
| **Pre-time-zero Gastrointestinal issues** |  |  | 0.023 | 0.055 |
| No | 48758 (93.2%) | 977 (92.6%) |  |  |
| Yes | 3552 (6.8%) | 78 (7.4%) |  |  |
| **Pre-time-zero Low grade fever** |  |  | 0.039 | 0.067 |
| No | 51912 (99.2%) | 1043 (98.9%) | |  |
| Yes | 398 (0.8%) | 12 (1.1%) |  |  |
| **Pre-time-zero Myagia** |  |  | 0.023 | 0.050 |
| No | 49412 (94.5%) | 991 (93.9%) |  |  |
| Yes | 2898 (5.5%) | 64 (6.1%) |  |  |
| **Pre-time-zero Post-exertional malaise** |  |  | 0.046 | 0.063 |
| No | 49728 (95.1%) | 992 (94.0%) |  |  |
| Yes | 2582 (4.9%) | 63 (6.0%) |  |  |
| **Pre-time-zero Loss of Taste** | |  | 0.046 | 0.072 |
| No | 51946 (99.3%) | 1043 (98.9%) | |  |
| Yes | 364 (0.7%) | 12 (1.1%) |  |  |
| **Pre-time-zero Loss of Smell** | |  | 0.025 | 0.057 |
| No | 51935 (99.3%) | 1045 (99.1%) | |  |
| Yes | 375 (0.7%) | 10 (0.9%) |  |  |
| **Pre-time-zero Difficulty Sleep** |  |  | 0.041 | 0.065 |
| No | 47252 (90.3%) | 940 (89.1%) |  |  |
| Yes | 5058 (9.7%) | 115 (10.9%) |  |  |

### Supplementary Table 3: Selection Rules for 4–8-Month and 9–12-Month Follow-up Surveys Based on Trial Start Date and Available Visit Data

| **Trial time zero** | **surveys_4_8_months** | **surveys_9_12_months** |
| --- | --- | --- |
| **2021-01** | V7 (7 months; if not available, censor at V7) | V8, V9 (if both are available, use v9 data - 12 months; if only v8 is available, use v8 - 9months; if v8 is not available, censor at V8) |
| **2021-02** | V8 (8 months; if not available, censor at V8) | V9 (11 months; if not available, censor at V9) |
| **2021-03** | V8 (7 months; if not available, censor at V8) | V9 (10 months; if not available, censor at V9) |
| **2021-04** | V8 (6 months; if not available, censor at V8) | V9, V10 (if both are available, use v10 data - 12 months; if only v9 is available, use v9 - 9months; if v8 is not available, censor at V8) |
| **2021-05** | V8, V9 (If both V8 and V9 are available, use V9 - 8 months; If only V8 is available, use V8 - 4 months; If V8 is not available but V9 is, censor at V8.) | V10 (11 months; if not available, censor at V10) |
| **2021-06** | V9 (7 months; if not available, censor at V9) | V10 (10 months; if not available, censor at V10) |
| **2021-07** | V9 (6 months; if not available, censor at V9) | V10, V11 (if both are available, use v11 data - 12 months; if only v10 is available, use v10 - 9months; if v10 is not available, censor at V10) |
| **2021-08** | V9, V10 (if both are available, use V10 data – 8 months; if only V9 is available, use V9 – 5 months; if V9 is not available, censor at V9) | V11 (11 months; if not available, censor at V11) |
| **2021-09** | V9, V10 (if both are available, use V10 data – 7 months; if only V9 is available, use V9 – 4 months; if V9 is not available, censor at V9) | V11 (10 months; if not available, censor at V11) |
| **2021-10** | V10 (6 months; if not available, censor at V10) | V11 (9 months; if not available, censor at V11) |
| **2021-11** | V10, V11 (if both are available, use V11 data – 8 months; if only V10 is available, use V10 – 5 months; if V10 is not available, censor at V10) | V12 (12 months; if not available, censor at V12) |
| **2021-12** | V10, V11 (if both are available, use V11 data – 7 months; if only V10 is available, use V10 – 4 months; if V10 is not available, censor at V10) | V12 (11 months; if not available, censor at V12) |
| **2022-01** | V11 (6 months; if not available, censor at V11) | V12, V13 (if both are available, use v13 data - 12 months; if only v12 is available, use v12 - 10months; if v12 is not available, censor at V12) |
| **2022-02** | V11 (5 months; if not available, censor at V11) | V12, V13 (if both are available, use v13 data - 11 months; if only v12 is available, use v12 - 9months; if v12 is not available, censor at V12) |
| **2022-03** | V12 (8 months; if not available, censor at V12) | V13 (10 months; if not available, censor at V13) |
| **2022-04** | V12 (7 months; if not available, censor at V12) | V13 (9 months; if not available, censor at V13) |
| **2022-05** | V12, V13 (if both are available, use V13 data – 8 months; if only V12 is available, use V12 – 6 months; if V12 is not available, censor at V12) | V14 (12 months; if not available, censor at V14) |
| **2022-06** | V12, V13 (if both are available, use V13 data – 7 months; if only V12 is available, use V12 – 5 months; if V12 is not available, censor at V12) | V14 (11 months; if not available, censor at V14) |
| **2022-07** | V12, V13 (if both are available, use V13 data – 6 months; if only V12 is available, use V12 – 4 months; if V12 is not available, censor at V12) | V14, V15 (if both are available, use v15 data - 12 months; if only v14 is available, use v14 - 10 months; if v14 is not available, censor at V14) |
| **2022-08** | V13 (5 months; if not available, censored at v13) | V14, V15 (if both are available, use v15 data - 11 months; if only v14 is available, use v14 - 9 months; if v14 is not available, censor at V14) |
| **2022-09** | V14 (8 months; if not available, censored at v14) | V15 (10 months; if not available, censor at V15) |
| **2022-10** | V14 (7 months; if not available, censored at v14) | V15, V16 (if both are available, use v16 data - 12 months; if only v15 is available, use v15 - 9 months; if v15 is not available, censor at V15) |
| **2022-11** | V14, V15 (if both are available, use V15 data – 8 months; if only V14 is available, use V14 – 6 months; if V14 is not available, censor at V14) | V16 (11 months; if V16 is not available, censor at V16) |
| **2022-12** | V14, V15 (if both are available, use V15 data – 7 months; if only V14 is available, use V14 – 5 months; if V14 is not available, censor at V14) | V16, V17 (if both are available, use v17 data - 12 months; if only v16 is available, use v16 - 10 months; if v16 is not available, censor at V16) |

### Supplementary Table 4: Per-protocol analog estimates using IPTW and IPCW: weighted incidence, incidence difference, and incidence ratio (with 95% confidence intervals) of incident Long COVID symptoms at 4–8 and 9–12 months of follow-up. CHASING COVID Cohort, including CSTE infection (infections between January 2021 – December 2022 with follow-up through December 2023).

a) Overall long COVID symptoms

| **Follow up time** | **Exposure Arm** | **IPTW and IPCW-adjusted risk (95% CI)** | | **Risk difference (95% CI)** | | **Risk ratio (95% CI)** | |
| --- | --- | --- | --- | --- | --- | --- | --- |
| 4-8 months | Participants with SARS-CoV-2 infection | 0.225 | 0.202, 0.247 | 0.112 | 0.089, 0.134 | 1.986 | 1.783, 2.188 |
|  | Participants without SARS-CoV-2 infection | 0.113 | 0.111, 0.115 | ref | ref | ref | ref |
| 9-12 months | Participants with SARS-CoV-2 infection | 0.181 | 0.160, 0.203 | 0.058 | 0.036, 0.080 | 1.467 | 1.289, 1.646 |
|  | Participants without SARS-CoV-2 infection | 0.124 | 0.121, 0.126 | ref | ref | ref | ref |

b) Long COVID with autonomic dysfunction symptoms

| **Follow up time** | **Exposure Arm** | **IPTW and IPCW-adjusted risk (95% CI)** | | **Risk difference (95% CI)** | | **Risk ratio (95% CI)** | |
| --- | --- | --- | --- | --- | --- | --- | --- |
| 4-8 months | Participants with SARS-CoV-2 infection | 0.105 | 0.091, 0.120 | 0.043 | 0.027, 0.057 | 1.683 | 1.436, 1.919 |
|  | Participants without SARS-CoV-2 infection | 0.063 | 0.061, 0.064 | ref | ref | ref | ref |
| 9-12 months | Participants with SARS-CoV-2 infection | 0.093 | 0.078, 0.109 | 0.027 | 0.012, 0.044 | 1.415 | 1.185, 1.675 |
|  | Participants without SARS-CoV-2 infection | 0.066 | 0.064, 0.068 | ref | ref | ref | ref |

c) Long COVID with exercise intolerance symptoms

| **Follow up time** | **Exposure Arm** | **IPTW and IPCW-adjusted risk (95% CI)** | | **Risk difference (95% CI)** | | **Risk ratio (95% CI)** | |
| --- | --- | --- | --- | --- | --- | --- | --- |
| 4-8 months | Participants with SARS-CoV-2 infection | 0.142 | 0.124, 0.159 | 0.076 | 0.057, 0.094 | 2.152 | 1.867, 2.454 |
|  | Participants without SARS-CoV-2 infection | 0.066 | 0.064, 0.068 | ref | ref | ref | ref |
| 9-12 months | Participants with SARS-CoV-2 infection | 0.113 | 0.096, 0.128 | 0.040 | 0.023, 0.056 | 1.549 | 1.313, 1.779 |
|  | Participants without SARS-CoV-2 infection | 0.073 | 0.071, 0.075 | ref | ref | ref | ref |

d) Long COVID with neurologic symptoms

| **Follow up time** | **Exposure Arm** | **IPTW and IPCW-adjusted risk (95% CI)** | | **Risk difference (95% CI)** | | **Risk ratio (95% CI)** | |
| --- | --- | --- | --- | --- | --- | --- | --- |
| 4-8 months | Participants with SARS-CoV-2 infection | 0.141 | 0.124, 0.157 | 0.064 | 0.047, 0.080 | 1.845 | 1.613, 2.056 |
|  | Participants without SARS-CoV-2 infection | 0.076 | 0.075, 0.078 | ref | ref | ref | ref |
| 9-12 months | Participants with SARS-CoV-2 infection | 0.117 | 0.101, 0.133 | 0.035 | 0.019, 0.052 | 1.435 | 1.236, 1.649 |
|  | Participants without SARS-CoV-2 infection | 0.081 | 0.079, 0.083 | ref | ref | ref | ref |

### Supplementary Figure 1: Heatmap of Individual-Level Weighted Symptom Count Over Four Time Intervals, Stratified by Symptom Cluster and COVID Status

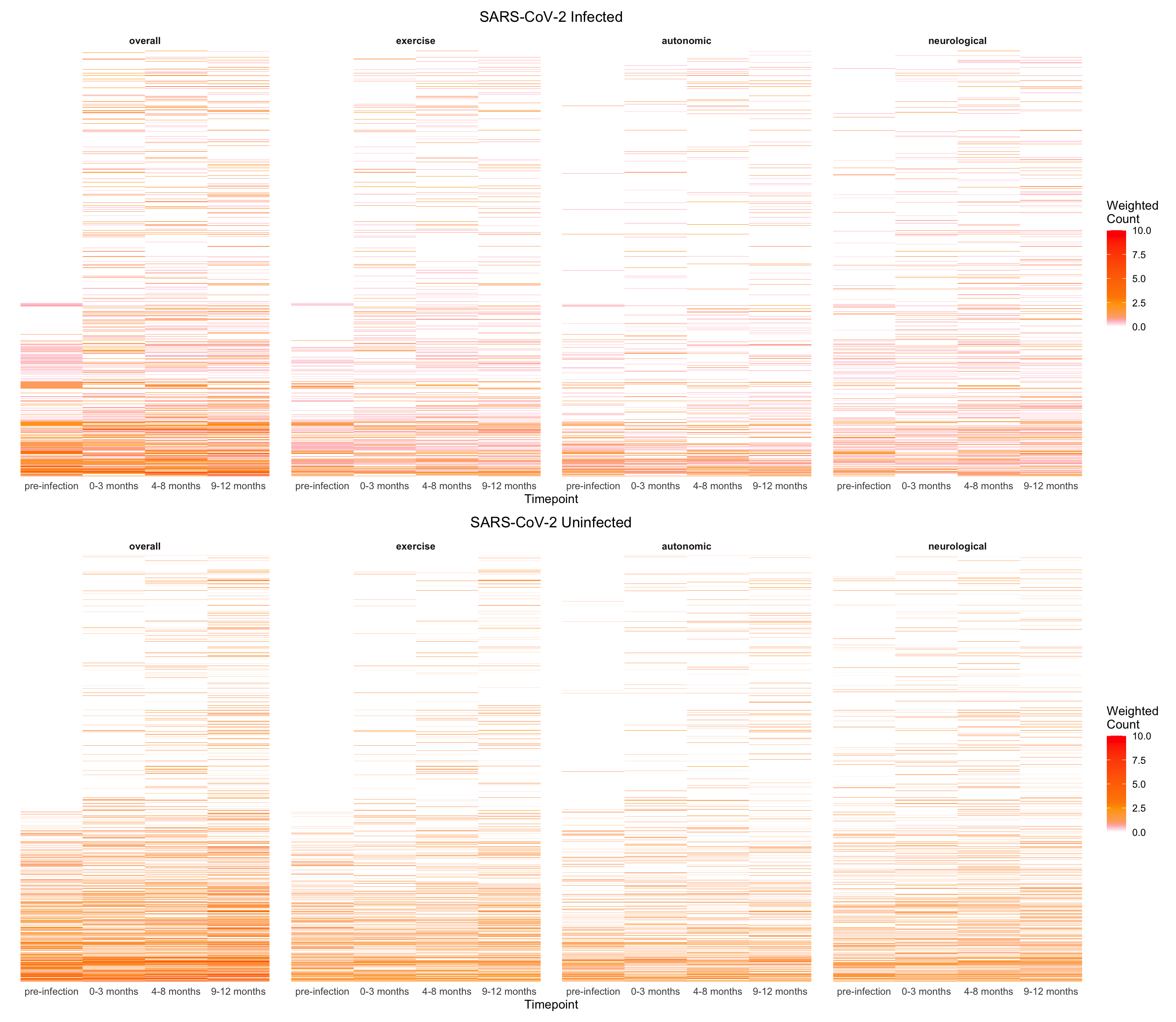

Supplementary Figure 2: Heatmap of Individual-Level Symptom Count Over Four Time Intervals, Stratified by Symptom Cluster and COVID/Long-COVID Status

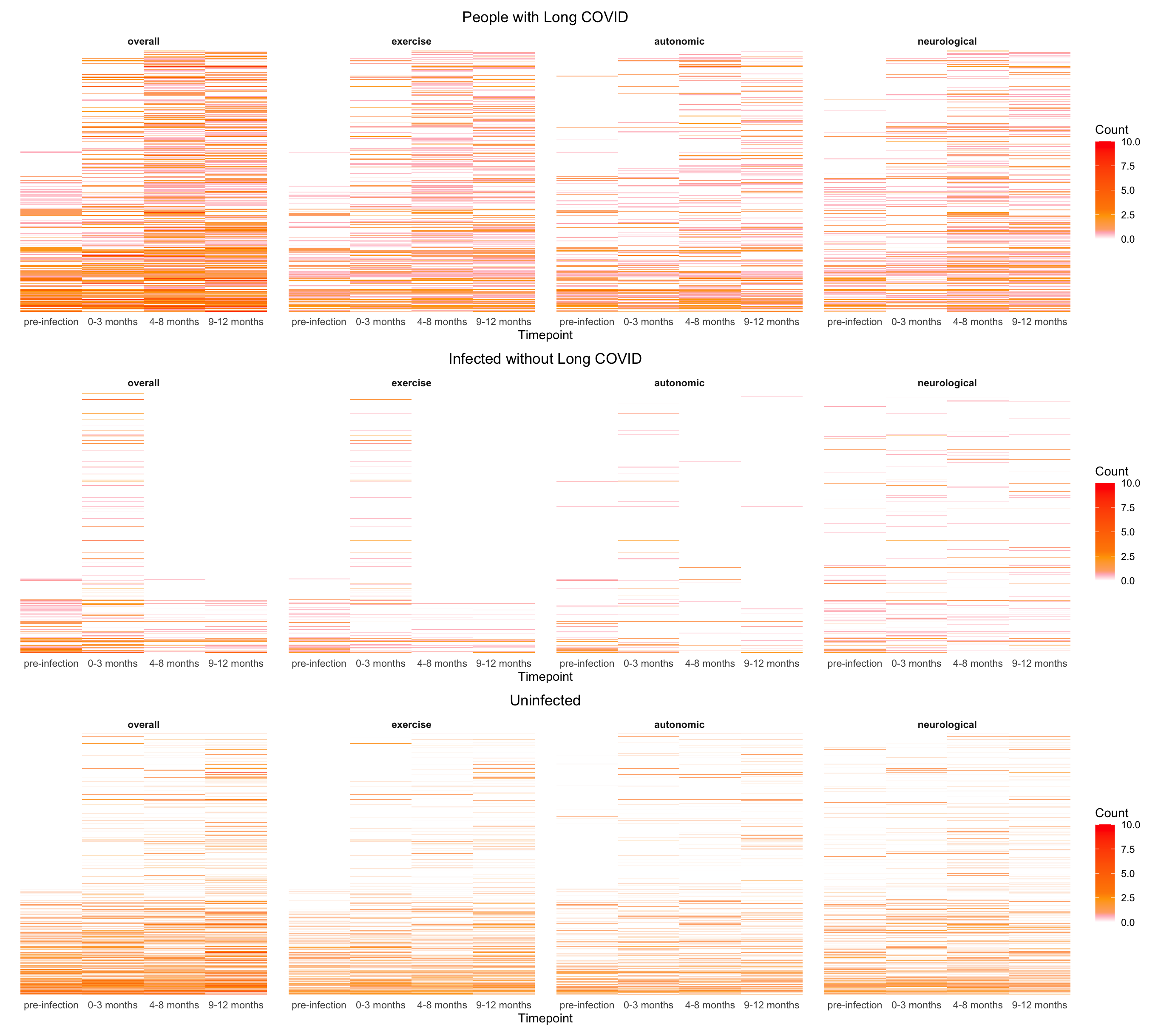

### Supplementary Figure 3: Flow Diagram of Inclusion Criteria for the CHASING COVID Cohort Study Sample for the First Sequential Trial

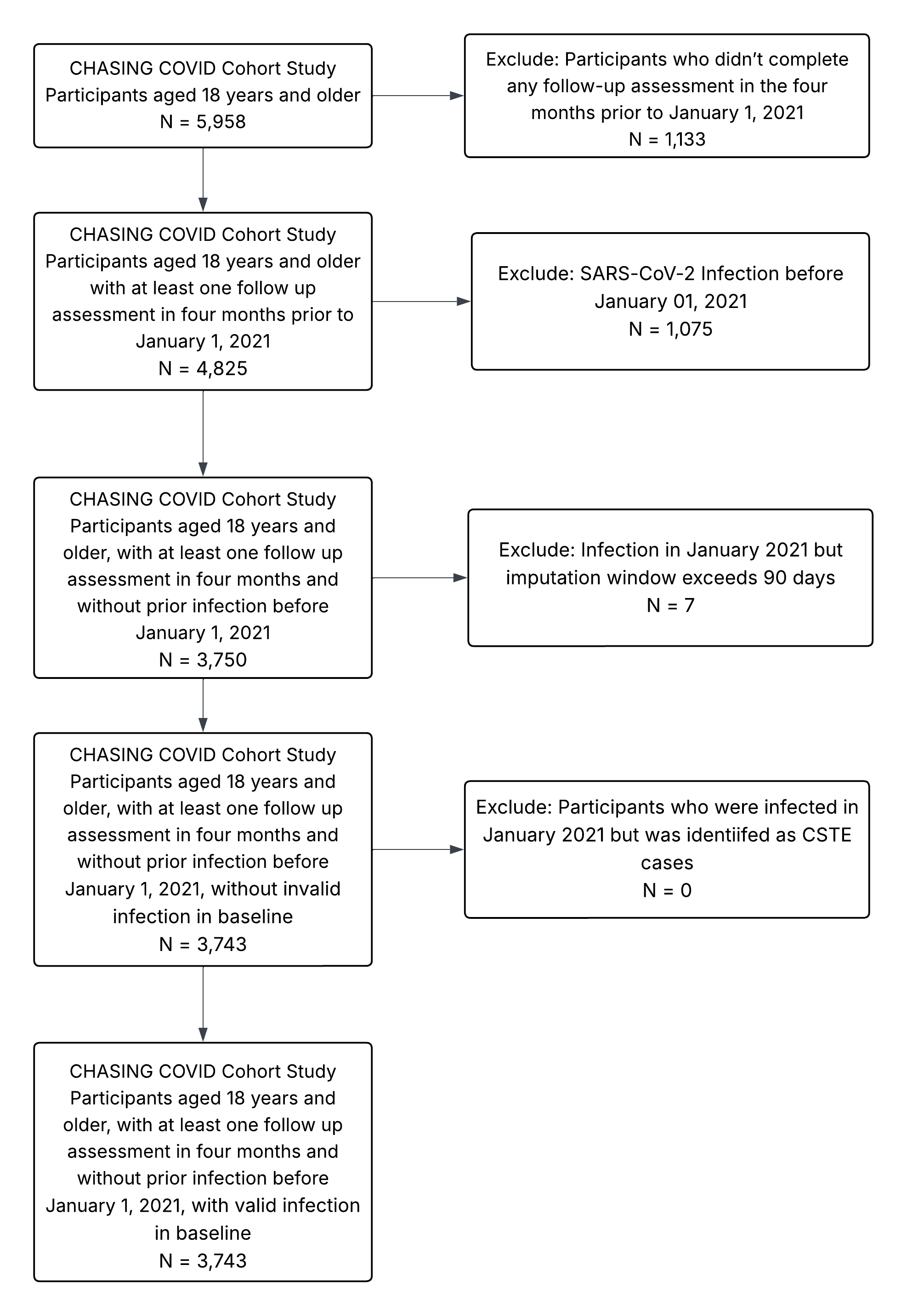
